# The association between *Helicobacter pylori* infection and risk of stroke: a comparison study between cross-sectional and longitudinal assessment of a 30-year follow-up

**DOI:** 10.1101/2025.09.12.25335672

**Authors:** Amenah Qotineh, Cyrille Kouambo, Queeneth Edwards, Susmita Dey, Shobhan Das, Olawumi Olatunde, Kelly L Sullivan, Logan Cowan, Jian Zhang

## Abstract

**Background:** Variations in study settings, designs, and background prevalence of *H. pylori* may contribute to the inconsistency regarding the relationship between *H. pylori* infection and stroke. To identify the causes of these inconsistencies and gain a deeper understanding of the potential link between *H. pylori* infection and stroke, we conducted this comparative study to assess the association from both cross-sectional and longitudinal perspectives using a single 30-year follow-up cohort with national representativeness.

**Method:** We analyzed data from 6,494 adults aged over 20 who participated in Phase I of the Third National Health and Nutrition Examination Survey (1988-1991) who were followed through December 31, 2019. We estimated adjusted hazard ratios (aHRs) of stroke mortality among participants who tested positive for *H. pylori* IgG antibodies compared to those who tested negative. Covariates included age, sex, race, family income, residence type, social isolation, alcohol consumption, serum cotinine, and job type.

**Results:** At baseline, adults who tested positive (n=3,141, 32.18%) had higher odds of being prevalent stroke cases [1.10 (1.04 - 1.15)] compared to non-infected adults. After 30 years of follow-up, among prevalent stroke cases (n=171), the aHRs of stroke deaths for *H. pylori*-positive participants were 1.54 (1.42 - 1.67), 1.64 (1.50 - 1.80), and 1.00 (0.74 - 1.36) for data ending in 2011, 2015, and 2019, respectively. The aHRs for all-cause deaths were 2.01 (1.71 - 2.36), 2.06 (1.74 - 2.43), and 2.01 (1.68 - 2.40). Among those who did not have cardio-cerebrovascular disease (CVD) history at baseline (n=5,969), the corresponding aHRs of stroke deaths for *H. pylori*-positive participants were 0.48 (0.34 - 0.68), 0.49 (0.34 - 0.69), and 0.59 (0.41 - 0.87) at the three time points.

**Conclusion:** (1) *H. pylori* infection was significantly associated with a history of stroke in cross-sectional analyses at baseline and increased the risk of dying from stroke, cancer, and all-cause mortality in these patients in prospective assessment over a 30-year follow-up period. However, among adults without a history of CVD, *H. pylori* infection was significantly associated with a reduced risk of stroke deaths. The protective effect of infection is manifested among alcohol drinkers and tobacco smokers only.

## Introduction

With the total cost approaching $50 billion, stroke remains a leading cause of death and disability in the US.^1^ Prevention efforts targeting well-defined, modifiable risk factors are crucial to addressing the growing stroke-related disease burden. In the past two decades, new evidence has emerged linking gut health and cardio-cerebrovascular disease (CVD), with some implicating *Helicobacter pylori (H. pylori)* infection in the development of stroke.^2^ *H. pylori* infection is relatively common, affecting about 30% to 40% of the US population ^3^ and about half of the global population.^4,5^ The treatment aiming to eradicate *H. pylori* may represent an impactful strategy for reducing the risk of stroke if the association is confirmed as causal.

Early studies based on small sample sizes, using case-control study designs in clinical settings, reported a relatively consistent association between *H pylori* infection and a low prevalence of stroke.^6–17^ Antibody levels to *H. pylori* predicted incident stroke.^18^ The seroprevalence of *H. pylori* was higher in prevalent stroke cases than in controls, and *H. pylori* eradication treatment was associated with a lower risk of stroke.^19,20^ However, longitudinal studies have reported inconsistent results. Some observed a significant association between *H. pylori* seropositivity and the onset of carotid plaque ^21,22^, carotid atherosclerosis,^23,24^ and incident stroke, ^18,19,25^, while others did not. ^26–30^ Protective effects were also reported from cagA-positive *H*. *pylori* infection and fatal stroke. ^28^ ^29^

The variations in the studies’ settings, design, power, and background prevalence of *H. pylori* may contribute to the inconsistencies. Existing literature is mainly based on case-control^6–17^ or cross-sectional designs,^31^ ^32^, which simultaneously measured the exposure and outcome and likely introduced reverse causality.^33^ The existing studies were conducted across socioeconomic contexts, with most from developed economies^6,18,25,28,31,34^ and a few from developing countries, including Iran,^35^ China,^23^ ^20^, and Congo.^20^ To identify the causes of the inconsistency and gain a deeper understanding of the potential link between *H. pylori* infection and stroke, we conducted this comparison study to assess the association cross-sectionally and longitudinally within a single prospective cohort followed for 30 years. We anticipate that the association between *H. pylori* and stroke may vary between the cross-sectional and longitudinal assessments within the same study population.

## Methods

### Study population

For the cross-sectional part of our study, the NHANES is a cross-sectional national survey sampling of the U.S. non-institutionalized population to assess the health and nutritional status of the general population. We restricted the analyses to 6,494 adults 20 years or older who participated in phase I of NHANES III, were eligible for follow-up (the second component of our study), and had non-equivocal results of the *H. pylori* test. After excluding participants who did not have data on family income, social isolation, and other key variables, a sample size of 6,494 was retained for analysis (**Figure 1**). A unique feature of NHANES is that the sampling approaches, interviews, and examination methods are standardized across surveys.^36^ The NHANES III started in 1988 and ended in 1994. The NHANES protocol was reviewed and approved by the National Center for Health Statistics’ Institutional Review Board (IRB), and the current study was exempt from ethics review by the IRB of the authors’ institution.

**Figure 1.** Flow Chart of the Study Population, 3 116 US Adults Aged 20 and Older,^1^ NHANES 1988 – 2019 **NDI=National Death Index; NHANES=the National Health Examination and Nutrition Survey**

### Baseline Data Collection – the platform for cross-sectional analyses

Baseline data were collected as part of NHANES III. Demographic and health-related information, including data on prescription medications, dietary intake, and food supplements, were gathered using standardized questionnaires. Under controlled and consistent environmental conditions, trained technicians drew blood and processed other biochemistry specimens in the Mobile Examination Centers for transportation to appropriate laboratories.^36^

#### Assessment of H. pylori infection

Stored serum samples were available for 6,494 participants aged 20 years and older who were interviewed and examined during phase 1 of NHANES III. *H. pylori* seropositivity was determined using a commercially available enzyme-linked immunosorbent assay from Wampole Laboratories (now Inverness Medical), Princeton, New Jersey. The immune status ratio (ISR) was calculated by dividing the specimen’s optical density by the mean optical density of the three cutoff controls. Specimens were classified as negative if the ISR was 0– 0.90, equivocal if the ISR was 0.91–1.09, and positive if the ISR was ≥ 1.10. Participants with equivocal results (n = 97, 2.5%) were excluded from the current analyses to minimize misclassification. This assay was tested against samples of 268 biopsy-confirmed *H. pylori* patients and 105 patients without *H. pylori* revealed a sensitivity of 91% and a specificity of 96%.^37^

#### Identification of prevalent stroke

In the NHANES interview, the participants were asked, "Has a doctor or other health professional ever told you that you had any of the following conditions?" This was followed by a list of various chronic and acute conditions representing prevalent conditions in the US population, including asthma, arthritis, cancer, chronic bronchitis, diabetes, hypertension, gout, lupus, stroke, heart attack, congestive heart failure, and thyroid disease. The responses to this question were used to assess the prevalence of stroke and other chronic conditions.^37^

#### Other covariates

Ethnicity was coded as white, black, or Mexican-American, with all remaining ethnicities categorized as ‘Other’. Family income level was assessed using the poverty income ratio (PIR), which was calculated using the previous year’s family income, and the family size was compared to the midpoint for the category and the family size with the federal poverty line (PIR=1). Respondents self-reported drinking, which was categorized as ‘rare/never’ or ‘any amount of drink. Self-reported cigarette smoking was classified as “rare/never,” “former,” and “heavy smokers” if they reported smoking 40 or more cigarettes in the past five days and as “moderate/light smokers” if they smoked fewer than 40 but more than zero cigarettes. Serum cotinine was measured to control for recent exposure to nicotine from tobacco. Dietary intake of folic acid was estimated using a 24-hour dietary recall. Prescription drug data were obtained by trained interviewers who inventoried all prescription drugs used within the past month. To prevent over-adjustment, the Healthy Eating Index (HEI) was used to indicate overall diet quality in the supplemental analyses to control for confounding from other nutrients and fiber. HEI measures the degree to which diets comply with specific recommendations in the Dietary Guidelines and the Food Guide Pyramid ^38^

### Vital status follow-up and identification of stroke death – the endpoints for longitudinal analyses

Twelve identifiers (including social security number, sex, and date of birth) were used to link NHANES III participants with the National Death Index to ascertain vital status and cause of death. The cause of death was determined using the underlying cause listed on the death certificate. The International Classification of Diseases, Injuries, and Causes of Death (ICD) 10th revision was used to code the deaths into several main groups by cause. All deaths before 1999 coded under ICD-9 were recorded into the comparable ICD-10 code. Three data cut points were used to assess the regression dilution effects: December 31, 2011; December 31, 2015; and December 31, 2019. The person-year contribution from each participant was calculated as the time between the baseline examination and the date of death (if it occurred) or the date of data cutoff (if still alive), whichever occurred first.

### Statistical analysis

We used SAS (SAS 9.4, Research Triangle Park, NC) procedures for surveys ^39^ and divided the analyses into three components **(Figure 1**):

(1) A cross-sectional assessment of the association between prevalent stroke and *H. pylori* seroprevalence was conducted with all participants older than 20 years who had data on both *H. pylori* test results and the history of stroke at baseline.
(2) A longitudinal assessment of the association between the risk of dying from major chronic diseases (diabetes, cancer, stroke, and heart disease) and *H. pylori* infection was performed among patients with prevalent stroke at the baseline survey.
(3) A longitudinal assessment of the association between the risk of dying from stroke and *H.* pylori infection was conducted on participants who did not have a CVD history at baseline.

For each component, we used the group with *H. pylori* antibody tested negative as the reference group and estimated the odds ratios (ORs – component 1) of being prevalent stroke or hazard ratios (HRs – component 2 and 3) of dying from stroke for the group tested positive adjusted for selected covariates. The proportional hazards assumption was checked by assessing the p-value of the interaction term between *H. pylori* infection and survival time. For component (3), we also performed additional analyses stratified by cigarette smoking, alcohol drinking, and duration of follow-up. Sensitivity analyses were conducted to evaluate the robustness of the estimated associations. The association between *H. pylori* infection and death from stroke or other chronic diseases could be confounded by well-established associations between CVD and dietary fibers, minerals, other vitamins, and overall dietary quality. However, controlling these nutrition components may over-adjust the association. Therefore, we included these as part of a sensitivity analysis. A two-sided p-value less than 0.05 was considered significant.

## RESULTS

The mean age of NHANES participants included in the current analyses was 43.35 years (standard error = 0.16), with 8.45% of participants aged 70 or older at the baseline interview. Reflecting the general population at the time of the baseline survey, 11.82% (0.30%) of adults were living below the poverty line. Whites comprised approximately 80% of the participants, while Mexican-Americans accounted for 4.58%, mirroring the racial and ethnic demographics of the adult population in the late 1980s (**Table 1**). With 171 prevalent stroke cases identified at baseline, the prevalence of stroke was 1.51% (0.04%). The mean age of prevalent stroke cases was at 69.25 years, with half being 70 years or older. The seroprevalence of *H. pylori* infection was significantly higher in prevalent stroke cases compared to the general population, 51.82% versus 32.18%.

**Table 1.** The characteristics of the study participants included in different analytic components Adults aged 20 +, the NHANES III, the baseline survey 1988-1991^1^.

|  |  | All participants<br>(N=6,494) |  | CVD free participants<br>(n=5,969) |  | Prevalent stroke cases<br>(n=171) |  |
| --- | --- | --- | --- | --- | --- | --- | --- |
|  | level | n | %(se) <sup>2</sup> | n | %(se) <sup>2</sup> | n | %(se) <sup>2</sup> |
| **Age(mean, year)** | Continuous(mean) | 6,494 | 43.35 (0.16) | 5,969 | 42.34 (0.16) | 171 | 67.25 (0.15) |
| **Sex** | Woman | 3,203 | 51.89 (0.22) | 2,986 | 52.39 (0.27) | 85 | 53.11 (0.93) |
| **Age group** | 19-35 | 2,042 | 36.49 (0.26) | 2,031 | 38.20 (0.29) | 1 | 0.47 (0.01) |
|  | 35-44 | 1,193 | 22.87 (0.44) | 1,171 | 23.64 (0.46) | 3 | 3.02 (0.10) |
|  | 45-54 | 835 | 14.63 (0.13) | 787 | 14.63 (0.14) | 13 | 13.96 (0.92) |
|  | 55-69 | 1,277 | 17.56 (0.22) | 1,119 | 16.60 (0.24) | 47 | 33.55 (0.97) |
|  | 70+ | 1,147 | 8.45 (0.20) | 861 | 6.94 (0.18) | 107 | 48.99 (0.72) |
| **Family income level <sup>3</sup>** | Middle and high | 3,384 | 67.64 (0.29) | 3,161 | 68.40 (0.28) | 63 | 44.89 (0.74) |
|  | Near poor | 1,761 | 20.54 (0.21) | 1,575 | 20.09 (0.21) | 73 | 34.07 (0.63) |
|  | Poor | 1,349 | 11.82 (0.30) | 1,233 | 11.51 (0.26) | 35 | 21.04 (0.78) |
| **Race and Ethnicity** | Whites | 3,051 | 79.00 (0.55) | 2,739 | 78.85 (0.53) | 107 | 83.79 (0.89) |
|  | Blacks | 1,576 | 9.99 (0.28) | 1,465 | 9.92 (0.27) | 41 | 11.83 (0.55) |
|  | Mexicans | 1,657 | 4.58 (0.15) | 1,562 | 4.63 (0.17) | 19 | 1.86 (0.13) |
|  | Others | 210 | 6.43 (0.38) | 203 | 6.61 (0.35) | 4 | 2.53 (0.98) |
| **Type of resident area** | Metro area | 3,222 | 48.09 (0.44) | 2,995 | 48.35 (0.42) | 75 | 38.03 (1.01) |
|  | Others | 3,272 | 51.91 (0.44) | 2,974 | 51.65 (0.42) | 96 | 61.97 (1.01) |
| **Alcohol drinking** | Light/moderate | 2,032 | 36.08 (0.44) | 1,926 | 36.76 (0.47) | 28 | 13.32 (0.57) |
|  | Rare/never | 4,462 | 63.92 (0.44) | 4,043 | 63.24 (0.47) | 143 | 86.68 (0.57) |
| **Cigarette smoking** | Former | 1,713 | 25.66 (0.30) | 1,491 | 24.87 (0.29) | 69 | 39.20 (1.13) |
|  | Heavy | 804 | 17.47 (0.27) | 756 | 17.55 (0.28) | 16 | 14.34 (0.60) |
|  | Light/moderate | 1,008 | 13.70 (0.23) | 961 | 14.08 (0.25) | 14 | 5.14 (0.28) |
|  | Rare/never | 2,969 | 43.17 (0.43) | 2,761 | 43.49 (0.46) | 72 | 41.32 (0.96) |
| **H. pylori infection** | Positive | **3,141** | **32.18 (0.34)** | **2,815** | **31.20 (0.35)** | **107** | **51.82 (0.80)** |
| **Major chronic disease at baseline <sup>4</sup>** |  |  |  |  |  |  |  |
| **Diabetes** | Yes | 459 | 4.41 (0.08) | 344 | 3.63 (0.08) | 42 | 25.62 (0.49) |
| **Cancer** | Yes | 530 | 7.50 (0.25) | 423 | 6.85 (0.26) | 38 | 24.38 (0.30) |
| **Stroke** | Yes | 171 | 1.51 (0.04) | . |  | 171 | 100.0 (0.00) |
| **Heart disease** | Yes | 415 | 3.75 (0.08) | . |  | 61 | 35.82 (0.91) |

Note: CVD = Cardio-cerebral vascular diseases, SE=standard error; NHANES=the National Health Examination and Nutrition Survey.
1. The characteristics were measured at baseline conducted between the years 1988 to 1991. 2. Presented as a percentage (standard error) unless otherwise specified. The n was unweighted, but the % and its standard error were weighted. 3. Respondents were categorized based on Poverty Index Ratio (PIR), poor was defined as $PIR < 1.0$ ; near poor as $1 \leq PIR < 2$ , and middle to high income as $2 \leq PIR$ . 4. Multiple diseases could be reported by an individual.

The cross-sectional analyses revealed a significant association between *H. pylori* infection and prevalent stroke at baseline (OR = 1.10 [1.04 - 1.15], **Table 2**). This association appeared to be modified by major sociodemographic factors. *H. pylori* infection was associated with a higher prevalence of strokes in whites but a lower prevalence in blacks. Modifying effects were also observed in terms of family income level and marital status. Following up on 171 prevalent stroke cases identified at baseline for 30 years revealed that 147 of these cases died before December 31, 2011, due to causes other than strokes. The HR of all-cause mortality for *H. pylori*-positive cases compared to negative counterparts were 2.17 (1.84 - 2.56), 2.22 (1.88 - 2.62), and 2.16 (1.83 - 2.55) for data ending in 2011, 2015, and 2019, respectively. About 10% of the prevalent stroke cases died from stroke during the follow-up period, with an HR of dying from stroke being 1.98 (1.75 - 2.23), 2.01 (1.77 - 2.29), and 1.45 (1.31 - 1.61) at the three data cutoff dates. The risk of dying from cancer was also significantly higher among infected patients, with corresponding hazard ratios of 3.52 (1.17 - 10.6), 3.52 (1.17 - 10.6), and 3.16 (1.13 - 8.82). Further adjustments for BMI, endocrine, nutritional, and metabolic diseases, and immunity disorder medication did not meaningfully change the estimates.

**Table 2.**
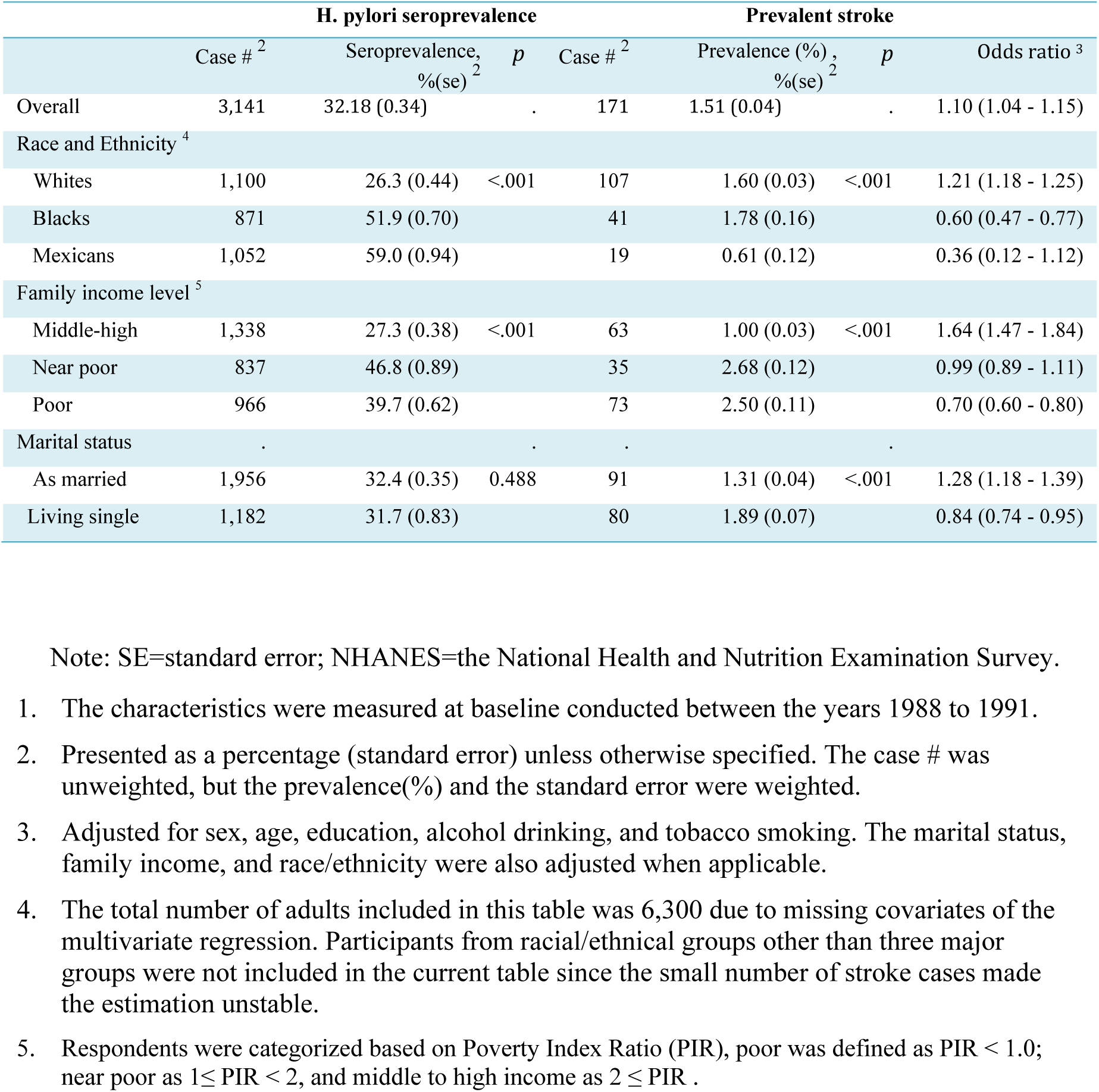
Stratified cross-sectional association between prevalent stroke and H. pylori infection at the baseline, 6,494 Adults aged 20 +, NHANES III 1988 – 1991^1^.

|  | H. pylori seroprevalence |  |  | Prevalent stroke |  |  |  |
| --- | --- | --- | --- | --- | --- | --- | --- |
|  | Case # <sup>2</sup> | Seroprevalence, % (se) <sup>2</sup> | p | Case # <sup>2</sup> | Prevalence (%) , % (se) <sup>2</sup> | p | Odds ratio <sup>3</sup> |
| Overall | 3,141 | 32.18 (0.34) | . | 171 | 1.51 (0.04) | . | 1.10 (1.04 - 1.15) |
| Race and Ethnicity <sup>4</sup> |  |  |  |  |  |  |  |
| Whites | 1,100 | 26.3 (0.44) | <.001 | 107 | 1.60 (0.03) | <.001 | 1.21 (1.18 - 1.25) |
| Blacks | 871 | 51.9 (0.70) |  | 41 | 1.78 (0.16) |  | 0.60 (0.47 - 0.77) |
| Mexicans | 1,052 | 59.0 (0.94) |  | 19 | 0.61 (0.12) |  | 0.36 (0.12 - 1.12) |
| Family income level <sup>5</sup> |  |  |  |  |  |  |  |
| Middle-high | 1,338 | 27.3 (0.38) | <.001 | 63 | 1.00 (0.03) | <.001 | 1.64 (1.47 - 1.84) |
| Near poor | 837 | 46.8 (0.89) |  | 35 | 2.68 (0.12) |  | 0.99 (0.89 - 1.11) |
| Poor | 966 | 39.7 (0.62) |  | 73 | 2.50 (0.11) |  | 0.70 (0.60 - 0.80) |
| Marital status |  |  |  |  |  |  |  |
| As married | 1,956 | 32.4 (0.35) | 0.488 | 91 | 1.31 (0.04) | <.001 | 1.28 (1.18 - 1.39) |
| Living single | 1,182 | 31.7 (0.83) |  | 80 | 1.89 (0.07) |  | 0.84 (0.74 - 0.95) |

Note: SE=standard error; NHANES=the National Health and Nutrition Examination Survey.
1. The characteristics were measured at baseline conducted between the years 1988 to 1991. 2. Presented as a percentage (standard error) unless otherwise specified. The case # was unweighted, but the prevalence(%) and the standard error were weighted. 3. Adjusted for sex, age, education, alcohol drinking, and tobacco smoking. The marital status, family income, and race/ethnicity were also adjusted when applicable. 4. The total number of adults included in this table was 6,300 due to missing covariates of the multivariate regression. Participants from racial/ethnic groups other than three major groups were not included in the current table since the small number of stroke cases made the estimation unstable. 5. Respondents were categorized based on Poverty Index Ratio (PIR), poor was defined as $PIR < 1.0$ ; near poor as $1 \leq PIR < 2$ , and middle to high income as $2 \leq PIR$ .

**Table 3.**
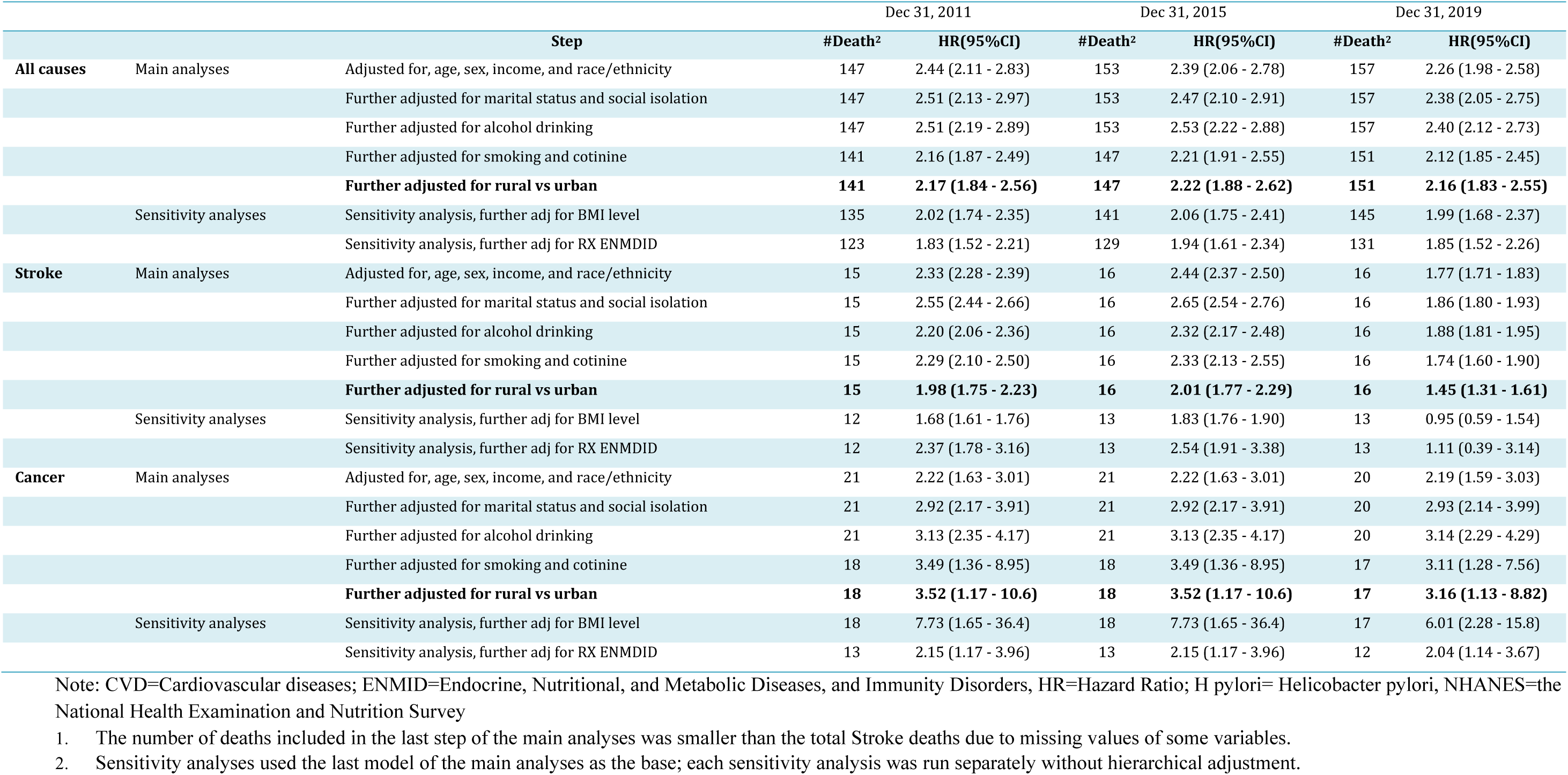
Hierarchically adjusted hazard risk ratios[HR(95%CI)] of major causes of death associated with Helicobacter pylori infection 171 prevalent stroke cases aged 20+, NHANES III follow-up study^1^.

After an average follow-up of 20 years (with a maximum of 31 years) and 141,410 total person-years (weighted person-years = 1,580 million person-years, S 1) of follow-up on 5,969 adults who were CVD-free at baseline, 157 stroke-related deaths were recorded, with 128 deaths occurring before the end of 2011. Stroke-related survival time was roughly one year longer for adults who tested negative compared to those who tested positive by the end of 2019, at 29.90 years vs. 28.58 years (**Figure 2**). However, after adjusting for age, infected individuals had a lower hazard of dying from stroke compared to those who were not infected. The hazard ratios (HRs) were 0.58 (0.38 – 0.84), 0.56 (0.37 – 0.84), and 0.68 (0.45 – 1.02) for data cut-offs on December 31, 2011, 2015, and 2019, respectively. Adjusting for additional covariates strengthened the association regardless of the follow-up length (**Figure 3** and S3). The HRs were 0.48 (0.34 – 0.68), 0.49 (0.34 – 0.69), and 0.59 (0.41 – 0.87) for the three data cut-off dates. The protective effect was more substantial among drinkers and smokers but disappeared among non-drinkers and non-smokers (S3).

**Figure 2.**
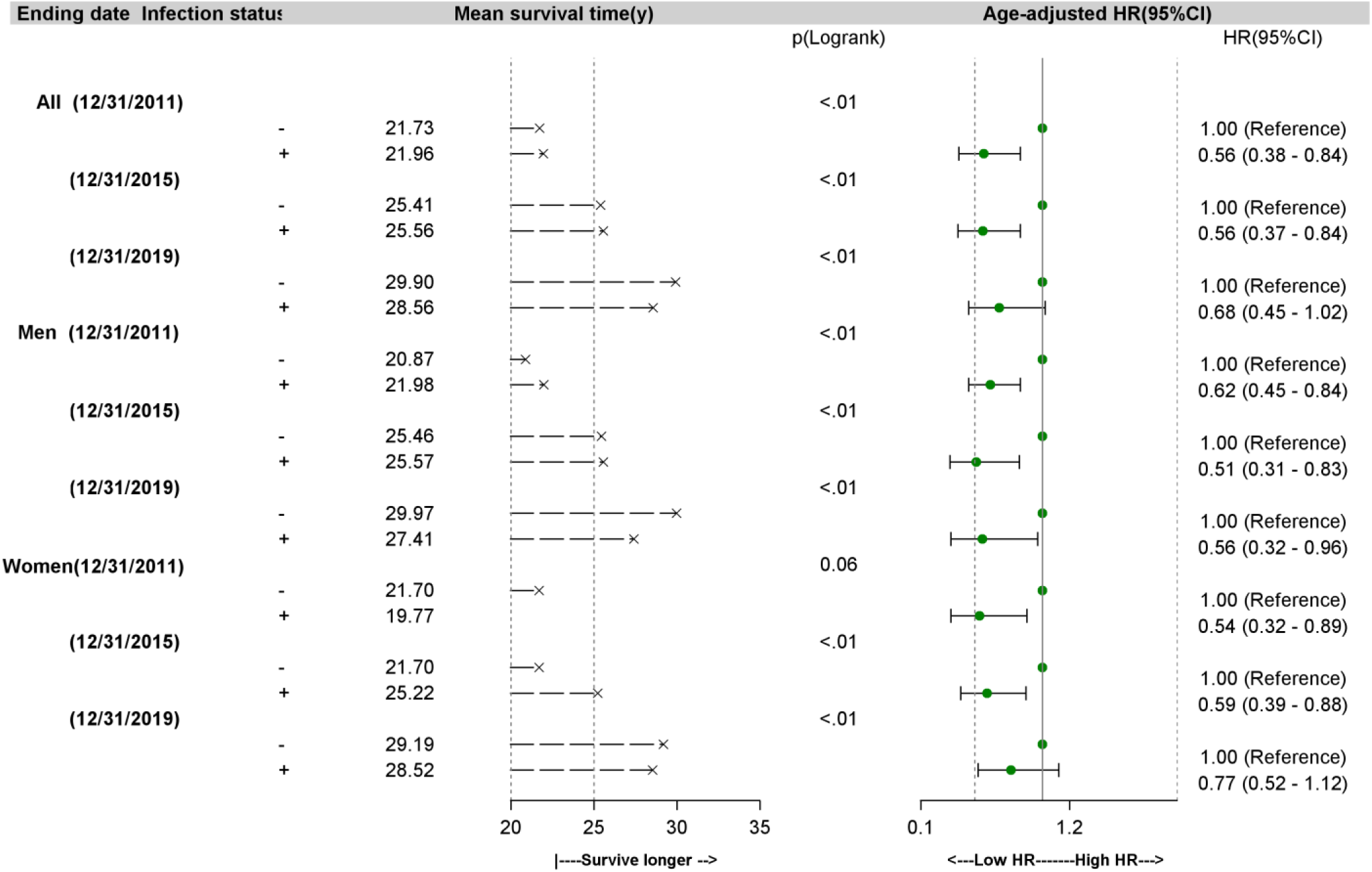
Survival time, mortality rate, and adjusted hazard risk ratio of stroke deaths associated with H. pylori infection 5,969 CVD-free adults aged 20+, NHANES III follow-up study 1988 - 2019 ^1,2 1^ Note: CVD=Cardio-cerebrovascular diseases; NHANES=the National Health Examination and Nutrition Survey. 1. The number of deaths included in the final model may be smaller than the total number of stroke deaths due to the missing value of some covariates. 2. P-value for Long-rank tests for the difference in survival year means between the study participants’ H pylori test positive and negative. 3. Per 1,000 person years. 4. Adjusted for age, sex, race, family income, alcohol drinking, cigarette smoking and serum cotinine, CVD history, vitamin B_12_ supplements. All were measured at the baseline survey.

**Figure 3:**
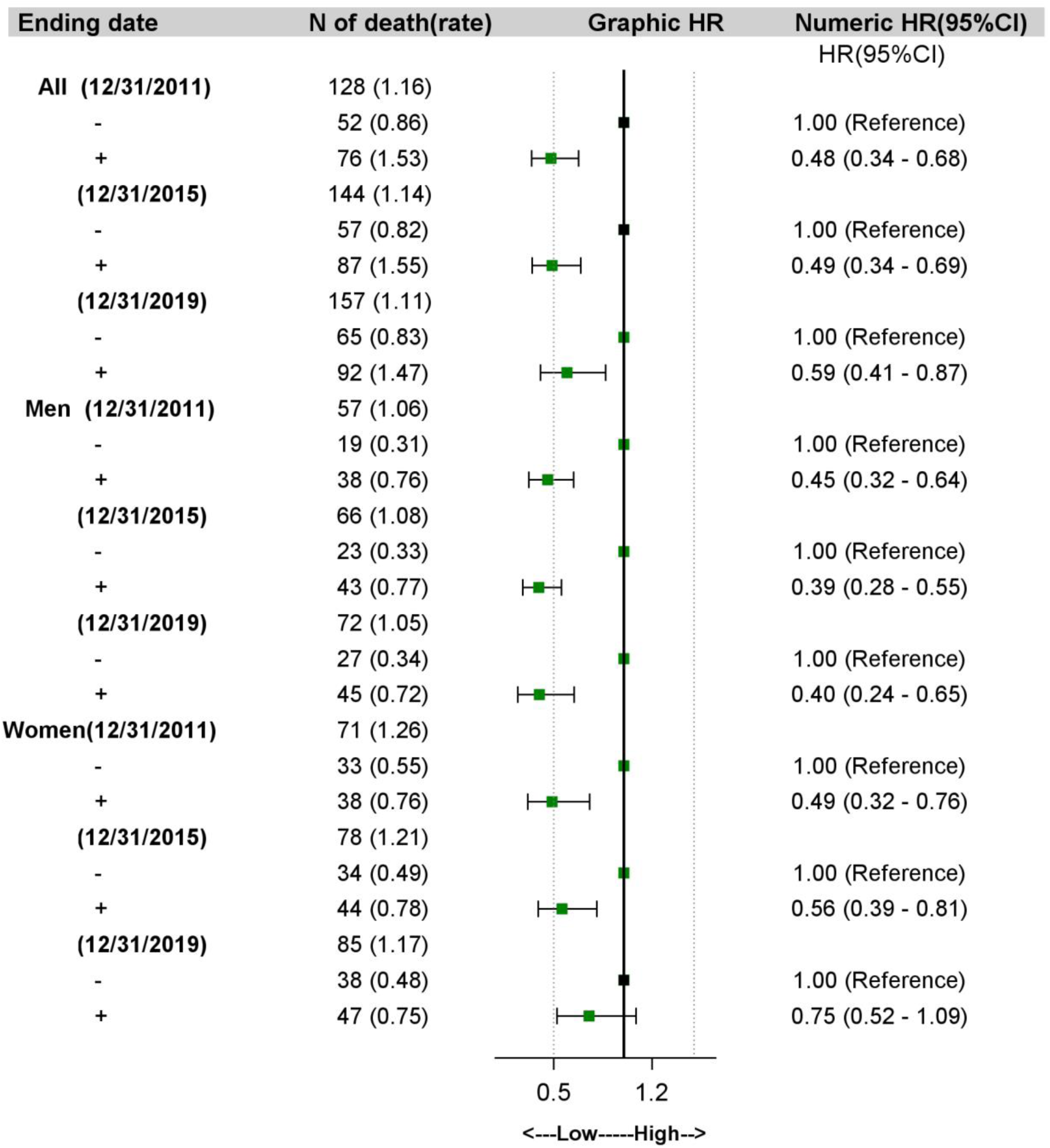
Adjusted hazard ratios for dying from Stroke associated with H. pylori infection 5,963 CVD-free adults aged 20+, NHANES III follow-up study 1988 - 2019 ^1,2^ Note: CI = Confident interval, HR = Hazard ratio; CVD=Cardio-cerebrovascular diseases; NHANES = the National Health Examination and Nutrition Survey. 1. The cutoffs for low and high can be found in the footnotes of the S1 2. The rate is expressed as per 1,000 person-year. 3. Adjusted for age, sex, race, family income, alcohol drinking, cigarette smoking, social isolation indicators (marital status, weekly frequency of visiting relatives, friends), serum cotinine, and type of jobs (white collars, blue collars, and services). All were measured at the baseline survey, 1988-1991.

## DISCUSSION

Using data rigorously collected in national surveys, we found a significant association between *H. pylori* infection and a history of stroke. After following up with these prevalent stroke cases, we observed that the risk of dying from stroke, cancer, and all-cause mortality was significantly elevated by *H. pylori* infection. However, the follow-up of those who were not diagnosed with any CVD at the baseline interview revealed that *H. pylori* infection was associated with a reduced risk of dying from stroke, and this protective effect persisted throughout the 30-year follow-up period.

The conclusion from the cross-sectional assessment in the current analyses was consistent with the literature. Using data from 147,936 individuals with a similar age range (25 years and older) and seroprevalence of *H. pylori* infection (52.0%) tested during 2002-2012, Shindler-Itskovitch et al. reported an adjusted OR of 1.16 (1.04, 1.29) and concluded that stroke may be associated with a history of *H. pylori* infection.^31^ Similarly, among 15,798 adults with *H. pylori* seroprevalence of 36%, *H. pylori* was associated with intracranial atherosclerosis in women.^32^ From a temporal perspective, case-control and cross-sectional studies share the same primary weakness, i.e., exposure and outcome are assessed simultaneously. The conclusion from the cross-sectional assessment in the current analyses aligns well with relevant case-control studies.^6–17^ However, further efforts are needed to clarify the modifying effects of sociodemographic factors on the relationship between *H. pylori* and prevalent stroke.

Following up with prevalent stroke cases for three decades, we observed that, consistent with the association obtained in the cross-sectional analyses, being infected with *H. pylori* at baseline significantly increased the risk of dying from stroke, cancer, and all-cause mortality. Stroke patients often require long-term use of anti-inflammatory medications, which can increase the risk of gastrointestinal bleeding.^40^ Additionally, stroke-induced immunosuppression can make patients more vulnerable to infections,^41^ complicating case management among stroke patients.<u>^42^</u>Conversely, inflammation initiated by *H. pylori* or other infectious pathogens may exacerbate the complications of a stroke, elevating the risk of dying from recurrent stroke or other causes.^42^ The proton pump inhibitors (PPIs), commonly used as part of the treatment regimen for eradicating *H. pylori infection*,^43,44^ should also be considered. Long-term use of PPIs was reported to cause morbidity, including cardiovascular disease.<u>^45^</u>Regardless of the direction of the association, taking consistent evidence from both case-control and cross-sectional studies, the current analysis, as the first one following prevalent stroke for more than 30 years, presents solid evidence to support the recommendations for managing *H. pylori* infection in patients with stroke or other CVDs.^43^

Following up with adults without a CVD history, we found that *H. pylori* infection was associated with a reduced risk of dying from stroke, and this protective effect persisted throughout the 30-year follow-up period. This report is not the first to reveal a protective effect from *H. pylori* infection in the general population after years of follow-up. Schottker et al. reported protective effects from *H. pylori* infection against fatal strokes. ^29^ Using the same data as the current analyses but with a shorter follow-up period, Chen added more evidence of a protective effect in NHANES participants. ^28^ The mechanism underlying the protective effect of *H. pylori* infection needs to be elucidated. It is prudent to consider that the infection may not be directly protective. Instead, eradicating *H. pylori*, likely occurring more among individuals who tested positive in the baseline survey, may reduce systemic inflammation and improve cardiovascular health. After more than 5 years of following up 4,765 adults with *H. pylori* infection but free of coronary heart disease, Kim et al. reported that *H. pylori* eradication prevented coronary heart disease, particularly in men ≤65 years and women >65 years.^46^ It was also suggested that a stroke might be more than just a break or blockage of cerebral blood vessels. Regulatory T-cells (T-reg cells), white blood cells that control the body’s immune response to prevent overreacting to harmful invaders, are postulated to be protective against stroke risk. In CVD-free adults with a T-reg down-regulated immune system, the damage to aging blood vessels in the brain might be less in *H. pylori-*positive hosts.^47,48^

It remains challenging to reconcile the protective effect of *H. pylori* infection among CVD-free adults and the elevated death risk associated with *H pylori* infection among stroke patients. The association between *H. pylori* infection and an increased risk of stroke or related deaths was reported to be substantially confounded by sociodemographic factors.^49^ ^35^ However, we adjusted for a set of identical sociodemographic factors. As a nonhomogeneous condition, the risk profile may differ between stroke subtypes. IgG antibody responses in *H. pylori* infection were found to be associated with an increased risk of stroke in the subtype of small-artery occlusion and a decreased risk of cardioembolic stroke. ^9,13,50^ Almost all gastric cancer deaths in the US are *H. pylori*-related;^51^ overestimation of the protective effect may occur due to survival bias or competing endpoints. The observation that the protective effects of infection manifested among alcohol drinkers and tobacco smokers only shines a light on the direction of future investigations.

The limitations of the current study are considerable. The small number of prevalent stroke cases prevented us from investigating more nuanced insights into the relationship. Although the study population was selected from a nationally representative sample, it excluded institutionalized groups where stroke and *H pylori* infection were more common. The validity of the death attributed to stroke in the certificate has been found to have a high specificity but a moderate sensitivity.^52,53^ The errors of the diagnoses in death certificates were not expected to be differential by *H. pylori* status, resulting in underestimations of the association rather than a spurious one. *H. pylori* infection was tested once. The “spontaneous” loss of the organism among those with established colonization was found to be low in earlier years.^54^ However, the widespread use of antimicrobial therapy against *H. pylori* began in the late 1980s,^44^ and *H. pylori* eradication treatment has become more common. On the other hand, acquiring *H. pylori* later in life is relatively common, especially among medically and socially disadvantaged populations.^44^ The bias associated with misclassification due to acquisitions or loss of *H. pylori* after baseline survey is unclear. Lumping all strokes together as one homogeneous entity is subject to criticism. The strengths of the current analyses are notable. The richness of NHANES data and the sustainably long follow-up period allow us to delineate the relationship with more extensive temporality after adjusting for a wide array of covariates. Compared with previous studies, the participants of NHANES III were selected from the community-dwelling population, extending the observations from clinical populations to general populations.

The association between *H. pylori* infection and stroke risk is complex and not fully understood. Although inconsistencies remain between studies following up with patients and those conducted among stroke-free populations, a more evident pattern is emerging. *H. pylori* infection appears to exacerbate inflammation and elevate the risk of dying from common chronic diseases in stroke survivors. Greater emphasis should be placed on *H. pylori* eradication in stroke case management. Further research is imperative to address the inconsistency between studies performed on patients and non-patients.

## Data Availability

The data that support the findings of this study are publicly available from the National Health and Nutrition Examination Survey (NHANES) conducted by the Centers for Disease Control and Prevention (CDC) at https://www.cdc.gov/nchs/nhanes/.

## Acknowledgments

The NHANES has been developed and funded by multiple federal agencies and is operated by the National Center for Health Statistics. The fieldwork was conducted by Westat, Inc. under contract to the National Center for Health Statistics. Conflict of Interest: This manuscript was initially prepared as a group project for graduate students enrolled in a seminar on chronic disease epidemiology led by Dr. Zhang at Jiann-Ping Hsu College of Public Health, Georgia Southern University. No funding was obtained to conduct the study, and all co-authors declare that they have no conflicts of interest.

## Author Contributions

JZ, LC, KLS designed research (project conception, development of overall research plan, and study oversight); AQ, CK, QE, SD, SD, OO conducted research and performed statistical analysis; NM and AQ, CK, QE, SD, SD, OO wrote the paper; AQ had primary responsibility for final content. All authors have read and approved the final manuscript.

